# Low-Cost Manually Assembled Open Source Reader for Isothermal Pathogen Detection from Saliva using RT-LAMP: SARS-CoV-2 Use Case

**DOI:** 10.1101/2020.10.19.20215319

**Authors:** John C. Bramley, Jason E. Waligorski, Colin L. Kremitzki, Mariel J. Liebeskind, Alex L. Yenkin, Xinyuan E. Xu, Matthew A. Lalli, Jane A. O’Halloran, Philip A. Mudd, Stacey L. House, Robi D. Mitra, Jeffrey D. Milbrandt, William J. Buchser

## Abstract

Distributed “Point-of-Care” or “at-Home” testing is an important component for a complete suite of testing solutions. This manuscript describes the construction and operation of a platform technology designed to meet this need. The ongoing COVID-19 pandemic will be used as the proof-of-concept for the efficacy and deployment of this platform. The technology outlined consists of a one-pot, reverse-transcription loop-mediated isothermal amplification (RT-LAMP) chemistry coupled with a low-cost and user-assembled reader using saliva as input. This platform is readily adapted to a wide range of pathogens due to the genetic basis of the reaction. A complete guide to the construction of the reader as well as the production of the reaction chemistry are provided here. Additionally, analytical limit of detection data and the results from saliva testing of SARS-CoV-2, are presented. The platform technology outlined here demonstrates a rapid, distributed, molecular point-of-care solution for pathogen detection using crude sample input.

## Introduction

The rapid spread of SARS-CoV-2 illuminated the stress a pandemic can put on traditional disease-testing paradigms and their associated global supply chains. The spread of the virus requires extensive testing to quickly identify and control outbreaks. This has led to testing shortages and delays in results reporting, and therefore a need for the development of more distributed and less supply-chain-limited testing solutions. This manuscript describes one such solution through the development of a pathogen-agnostic, distributed, rapid, saliva-based, molecular testing platform. This platform consists of an isothermal reaction evaluated real time using a heated fluorescence reader. This manuscript details the design, assembly, and operation of the reader as well as the formulation of the reaction chemistry and the exploration of both laboratory and clinical effectiveness in a SARS-CoV-2 use case. SARS-CoV-2 testing can be conducted using a wide range of techniques and technologies. A common gold standard is sample collection via nasopharyngeal swab followed by RNA isolation, then detection through RT-qPCR. However, shortages in reagents and swabs, as well as testing backlogs, have created the need for additional testing systems. Recent advances, by us and others, in saliva-based testing have expanded the tools available to centralized testing facilities and have the potential to reduce the supply chain burden of NP swab-based testing (Becker et al., 2020; M. A. A. Lalli et al., 2020). However, advancements for centralized lab facilities are not able to solely address the broad spectrum of testing needs under the challenge of a pandemic. Smaller healthcare facilities, as well as rural and remote areas, are not equipped to house these types of facilities and delays caused by sample transportation reduce the impact of testing. Thus, the need for rapid, *distributed*, and accessible “point-of-care” solutions. The system proposed here was designed specifically to provide a low-cost point-of-care testing solution using readily available components.

The development of this platform was guided by a core set of principles. *1 Distributed*. The instrumentation should not be reliant on a centralized laboratory facility and allow testing to be conducted outside of a CLIA environment. *2 Molecular (genetic)*. The advantages of this are two-fold, first is being *pathogen agnostic*, meaning primers can be replaced to identify other pathogens, and the second is the high specificity afforded by genetic-based tests. *3 “One-Pot” Chemistry*. No sample preprocessing is necessary to enhance ease of use and eliminate the need for microfluidics or cartridges. *4 Real time fluorescence*. We sought to avoid the problems of both colorimetric and lateral flow-based assays, namely the potential for ambiguity in results and interpretation. The advantages of a real-time fluorescent system are the ability to determine results analytically and to automatically report to a digital record or database.

The platform presented here will illustrate the design, construction, and performance of a rapid genetic test using SARS-CoV-2 as a proof of concept. This system consists of a low-cost, easy-to-assemble, fluorescent reader and one-pot, saliva-based, reverse-transcription loop-mediated, isothermal amplification (RT-LAMP) reaction chemistry. A full guide to the assembly and use of this testing platform is provided. Additionally, performance data in a simulated clinical matrix using viral particles as well as an experiment on clinical saliva samples is also reported.

### LAMP and Saliva Chemistry

The choice of assay for point-of-care testing must satisfy several criteria in order to meet the demands of the project. Traditional qRT-PCR requires sample pre-processing (RNA extraction) as well as thermal cycling, which increases the complexity of the instrumentation and handling of the sample. Isothermal reactions, therefore, provide a suitable alternative. Specifically, reverse-transcription loop-mediated isothermal amplification (RT-LAMP) has shown to be effective in viral detection using a crude sample as input (Augustine et al., 2020; Chotiwan et al., 2017; Das, Babiuk, & McIntosh, 2012; Kaneko, Kawana, Fukushima, & Suzutani, 2007). RT-LAMP provides a sensitive platform for nucleic acid detection in less than 30 minutes and is compatible with the reader described in this manuscript. Saliva-based testing was the preferred route for this platform due to the ease of collection, patient comfort, and testing worker safety. High viral loads have also been detected in saliva samples throughout the course of infection and at levels noticeably higher than nasopharyngeal swabs (La Scola et al., 2020; Wölfel et al., 2020). To overcome inhibitors present in saliva and create a test reaction that requires no sample processing (a “one pot reaction”), several modifications to the standard RT-LAMP reaction were needed (M. A. Lalli et al., 2020; Zhang et al., 2020). The modifications include the addition of 6 molar Guanidine hydrochloride, RNAsecure, 10% Tween 20, and additional BST 2.0 polymerase. The concentrations and protocol for creation of the master mix can be found in the methods section. This modified reaction overcomes the inhibitors found in saliva and is capable of robust nucleic acid detection in viral particles containing clinical matrix (saliva) as well as clinical samples.

### Volumetric Loop-Based Sample Collection

Commercial saliva collection devices are designed to hold several hundreds of microliters to milliliters in volume and preserve the samples for later assays, but these tubes (designed to minimize the exposure of the machine operator to infectious saliva) add complexity and cost to the assay. Since our point-of-care assay is performed immediately and requires only one microliter of saliva, we decided to implement volumetric inoculating loops for saliva collection (or self-collection). Similar saliva collection methods are currently in use in France and Israel for SARS-CoV-2 detection. Individually wrapped, sterile, plastic 1 microliter inoculating loops (EZ BioResearch K1021 and Cole-Parmer 06231-07) can be touched to the tongue then dipped directly into the LAMP reaction. This flexibility in sample collection reduces dependence on specialized manufacturing and diversifies the supply chain options as well as being extremely inexpensive (∼10x less than commercial saliva collection tubes).

## Methods

### Chemistry Production

#### Chemistry Production Reagent Preparation

A total of 12 primers (two sets of six) are used in the LAMP reaction chemistry. Each set targets a different genomic region, the E1 and N2 regions of the SARS-CoV-2 virus (Augustine et al., 2020; Yu et al., 2020). The purpose of the two sets is to increase sensitivity and add redundancy in the case of a partially degraded sample. All primers were ordered from Integrated DNA Technologies (IDT). The primer sequence information is listed in **Table 1**.

**Table 1.**
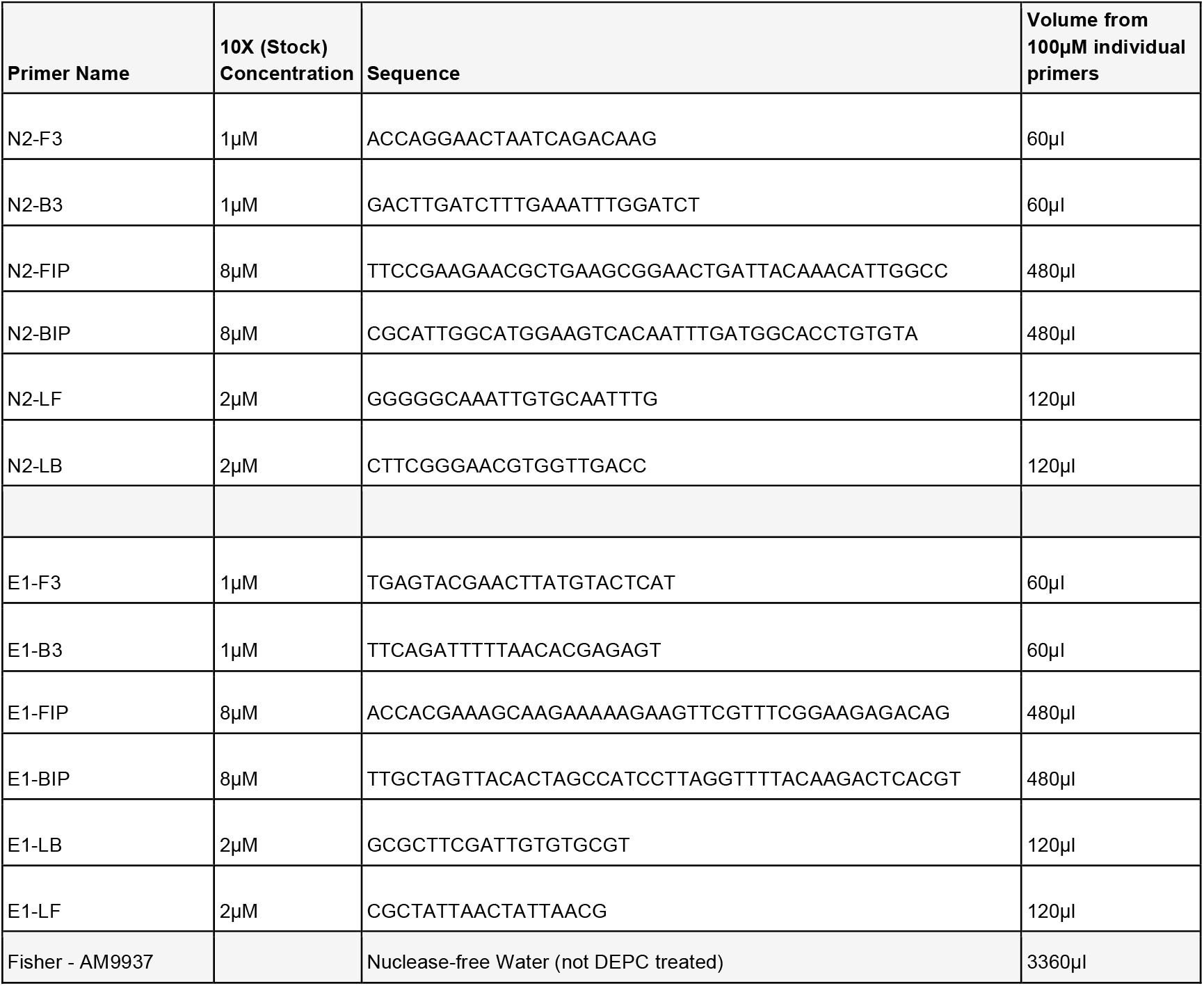
Primer sequences for the LAMP reaction. Two complete sets of LAMP primers are listed, one targeting the N2 region with the other targeting the E1 region. The starting point for each individual primer is 100µM, then they are combined to this 10x stock solution. The 10X primer concentration applies across both sets of primers reducing the concentration of each individual primer but maintaining the 10X concentration for all 12 primers.

Preparation of the LAMP reaction chemistry is done in a batch format and aliquoted into 0.2 mL PCR tubes (reaction tubes). The process described allows for the production of 432x 40µl reactions (40µl is the standard reaction volume used for testing). All reagent preparation should be done on ice and in a designated “Clean Space” free of DNA/RNA contaminants. The use of a PCR enclosure is advised to further minimize the risk of contamination. The LAMP enzyme and buffer, as well as fluorescent dye, are provided through a kit (NEB-E1700L), with additional enzyme (NEB – M0537L). Additionally, 6M Guanidine hydrochloride (EMD Millipore - G3272), 10% Tween 20 (EMD Millipore - P9416), and nuclease-free water (Fisher - AM9937) are also needed. All primers and gene block positive controls were acquired from Integrated DNA Technologies (IDT).

In order to prepare the chemistry, ensure that all reagents are thawed and the workspace has been cleaned with 10% bleach then 70% ethanol. Batches of reactions are prepared in 50mL RNAse-Free conical tubes prior to being aliquoted into 0.2mL PCR tubes. Add each reagent in the order found in **Table 2**. Ensure that reagents are mixed thoroughly through pipetting. Following reagent addition, the reactions can be aliquoted into the 0.2mL PCR tubes (40µl) and briefly centrifuged to eliminate any bubbles that may have formed. Mark ⅓ of the tubes with a blue dot designating them as negative controls leaving another ⅓ unmarked for sample testing. The remaining ⅓ will be used for positive controls.

**Table 2.** Chemistry reagent information.

| Volume to add in µl | Reagent |
| --- | --- |
| 5,076 | Nuclease-free Water (not DEPC treated) - (Fisher - AM9937) |
| 1,728 | 10X LAMP E1/N2 Primers (IDT, diluted in house) |
| 114.5 | 6M Guanidine hydrochloride (EMD Millipore - G3272, diluted in house) |
| 345.6 | RNAsecure (Fisher - AM7005) |
| 345.6 | Warmstart Dye (included in Warmstart LAMP Kit - NEB - E1700L) |
| 172.8 | 10% Tween 20 (EMD Millipore - P9416, diluted in house) |
| 8,640 | Warmstart Master Mix (Warmstart LAMP Kit - NEB - E1700L) |
| 864 | Bst 2.0 DNA Polymerase (NEB - M0537L) |

A positive control reaction is needed to ensure proper reader and reaction functionality. A non-biohazardous gene block is used in this reaction formulation. In the case of *positive control* reaction production, gene block needs to be added to the 50ml conical tube prior to aliquoting. Positive control template can be created by adding equal parts E1 gBlock (IDT - custom) and N2 control amplicon (IDT - 10006621) to a concentration of 10,000 parts per µl, diluted with 10:0.1mM TE buffer, pH 8.0. 144µl of diluted gene block should be added to the 50ml conical tube (equivalent to 1µl per 40µl reaction). Add 41µl of positive control containing reagents to 0.2mL PCR tubes and label with a red + to designate as a positive control. Reactions are stable at 4C for up to two days and several months at −20C.

E1 gBlock sequence 5’-3’:

GCCTTTGTAAGCACAAGCTGATGAGTACGAACTTATGTACTCATTCGTTTCGGAAGAGACAGGTACGTTAATAGTTAATAGCGTACTTCTTTTTCTTGCTTTCGTGGTATTCTTGCTAGTTACACTAGCCATCCTTACTGCGCTTCGATTGTGTGCGTACTGCTGCAATATTGTTAACGTGAGTCTTGTAAAACCTTCTTTTTACGTTTACTCTCGTGTTAAAAATCTGAATTCTTCTAGAGTTCCTGA

TC

10X E1/N2 LAMP Primer Recipe

LAMP Master Mix Recipe - 432 reactions

#### Reaction Chemistry Quality Control

In order to ensure that a given batch/lot of reaction chemistry is suitable for use, quality control measures are undertaken. Using a reader that has been assembled and quality controlled (outlined below), a minimum of four positive controls and four negative controls should be run. In each run, the negative control should show no amplification and the positive control should show amplification (X50 between 12 and 20 minutes). Negative controls showing more than a background amplification (X50 24 minutes) may be contaminated. It is unfortunately very easy to contaminate all your tubes with the positive control mix (for example from gloves > surface of pipette > new reaction tube), so take great care to separate the tubes, change gloves, and clean areas so that there is no possibility of positive-control contamination.

### Reader Construction

#### Electronics: Overview

We intended the reader to be constructed with few tools and readily available components. All components for the reader were acquired through consumer-accessible suppliers with several alternatives available. A circuit diagram of the reader can be found in **Figure 1**. The circuit consists of a series of six sensors powered by the 5-volt supply of the controller, two blue LED lights, buzzer, multicolor LED, and a 3.6-Watt heating element supplied by a 9-volt power supply. The 9-volt supply has been validated using both off-the-shelf 9V batteries and an AC power supply (at 0.55 Amps). The six sensors consist of photoresistors and thermistors that are housed within the “tube holder,” a 3-D printed part detailed below. An Arduino Nano with an ATMega chipset was selected as the reader’s controller. All product number information can be found in **Table 3**. The following electronics build can be most easily accomplished by referring to **Figure 2**. The construction of the reader requires a limited understanding of electronics, access to a 3D printer (or a printing service), and a small tool set consisting of wire strippers, forceps, hand-held saw, and scissors. A soldering iron can add robustness to elements of the construction and is recommended (but not required).

**Table 3.** Parts required. The first line of each row is the brand of the part and the second is the item name. The next column is an estimated price per single part (what we paid, but we often purchased in bulk). The 3rd column indicates how many of these go into each reader. The final column lists the manufacturer part number and the supplier part number.

| Component | ~Price | # per Device | Catalog # |
| --- | --- | --- | --- |
| Inland Nano<br>FT232 Set (contains main Board Nano) | \$14.99 | 1 | Mfr: CZ0350<br>Micro Center: 055251 |
| USB Connector | 0 | 1 | Included in Nano FT232 Set |
| Male-Male Jumper Ribbon | 0 | 8 | Included in Nano FT232 Set |
| Male-Female Jumper Ribbon | 0 | 6 | Included in Nano FT232 Set |
| LEDs (blue, red) | 0 | 2/1 | Included in Nano FT232 Set |
| Full Color RGB | 0 | 1 | Included in Nano FT232 Set |
| 10 K Ohm Resistors | 0 | 1 | Included in Nano FT232 Set |
| 470 Ohm Resistors | 0 | 3 | Included in Nano FT232 Set |
| 220 Ohm Resistors | 0 | 1 | Included in Nano FT232 Set |
| Bud Industries<br>170-Tie Point Breadboard | \$2.60 | 1 | Mfr: BB-32650-B<br>Digi-Key: 377-BB-32650-B-ND |
| Advanced Photonix<br>Photoresistors | \$0.54 | 2 | Mfr: PDV-P8104<br>Digi-Key: PDV-P8104-ND |
| On Semiconductor<br>NPN Transistors | \$0.09 | 1 | Mfr: KSP2222ABU<br>Digi-Key: KSP2222ABUFS-ND |
| Micro Commercial Co<br>PNP Transistors | \$0.09 | 2 | Mfr: PN2907A-AP<br>Digi-Key: PN2907A-APCT-ND - Cut Tape (CT) |
| Murata Electronics<br>Thermistors | \$0.34 | 2 | Mfr: NXRT15XV103FA1B040<br>Digi-Key: 490-7171-ND |
| Vishay Beyschlag/Draloric/BC<br>Components<br>7.5 Ohm Heating Resistors | \$0.05 | 3 | Mfr: PAC100007508FA1000<br>Digi-Key: PPC1D7.5CT-ND - Cut Tape (CT) |
| Stackpole Electronics Inc<br>10 K Ohm 1% Resistors | \$0.02 | 6 | Mfr: RNF14BTE10K0<br>Digi-Key: RNF14BTE10K0TR-ND - Tape & Reel (TR) |
| Stackpole Electronics Inc<br>100 Ohm 1% Resistors | \$0.10 | 1 | Mfr: RNMF14FTC100R<br>Digi-Key: S100CATR-ND - Tape & Reel (TR) |
| Keystone Electronics<br>Battery Clip | \$0.53 | 1 | Mfr: 235<br>Digi-Key: 36-235-ND |
| XP Power<br>AC/DC Wall Mount Adapter 9V 5W | \$6.50 | 1 | Mfr: VEL05US090-US-JA<br>Digi-Key: 1470-2773-ND |
| TDK Corporation<br>Buzzer | \$0.51 | 1 | Mfr: PS1240P02BT<br>Digi-Key: 445-2525-1-ND - Cut Tape (CT) |
| Adafruit Industries LLC<br>Heat Shrink | \$0.16 | 2 | Mfr:1649<br>Digi-Key: 1528-1649AD-ND |
| Adafruit Industries LLC<br>Wire (optional) | \$0.06 | 1 | Mfr: 2990<br>Digi-Key: 1528-1766-ND |
| Bud Industries<br>Wire Jumpers | \$0.07 | 1 | Mfr: BC-32626<br>Digi-Key: 377-2258-ND |
| 3D Printing Material, Printing Costs | \$5.00 | 4 | |
| Felt Insulation (about 2" square per reader) | \$0.03 | 2 | Any fabric store (Joann Fabric, Michaels) |
| Optical Filter<br>(We cut these from a larger piece that was used for UV protection goggles, but other sources are also likely available) | \$9.80 | 1 | Clare Chemical: AG16 |

**Figure 1.**
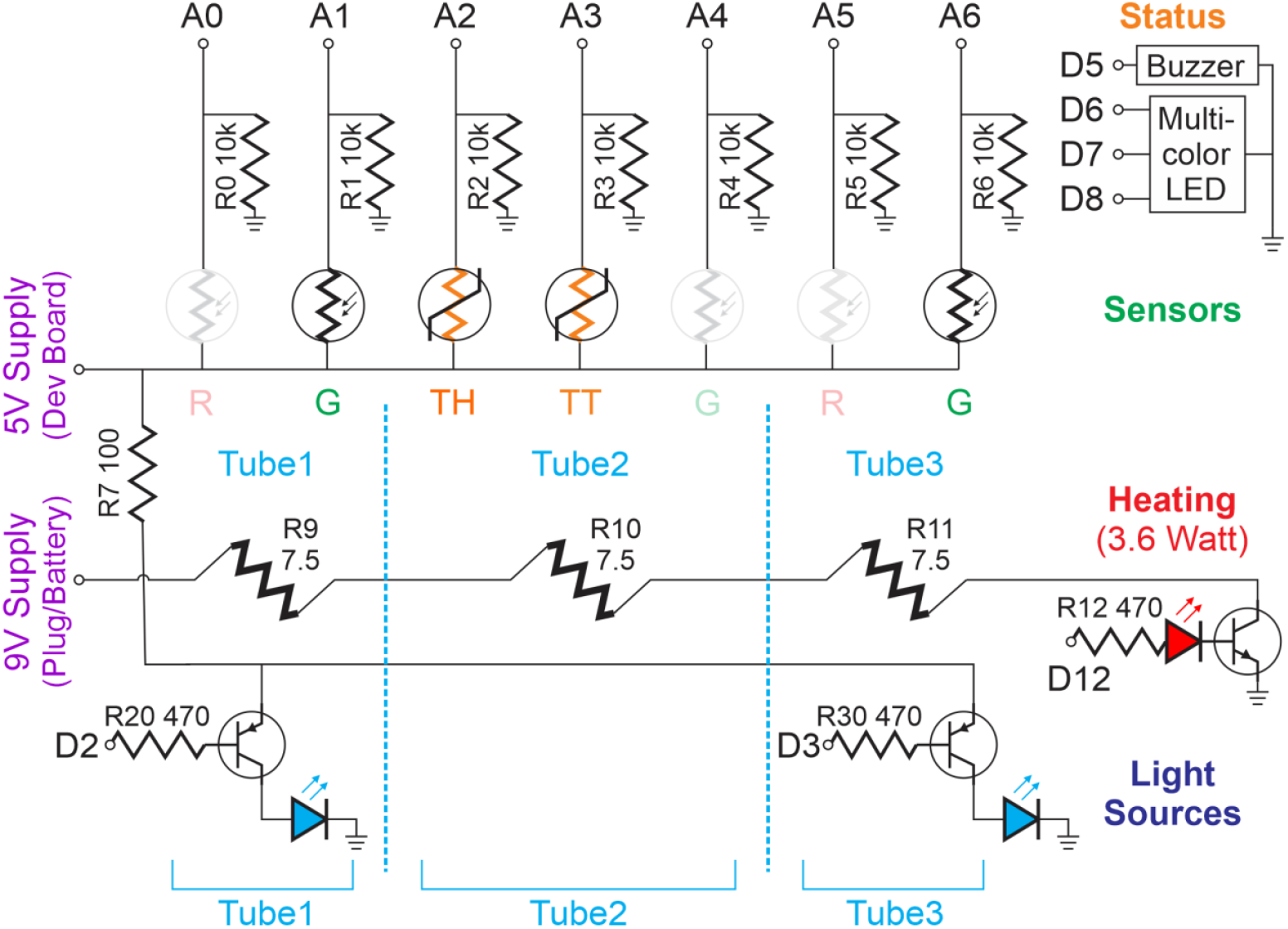
The circuit diagram. A0-A6 are the analog inputs and D2, D3, D5-D8, D12 are digital pins outputs of the Arduino nano. Note the transistor near R12 is an NPN BJT while the two transistors near R20 and R30 are PNP BJTs. The diagram shown allows for the system to be easily converted into a two-color fluorescence system (those elements are grayed out and are not necessary for the primary build in this paper). Each tube position is also indicated showing which components relate to which reaction tube.

**Figure 2.**
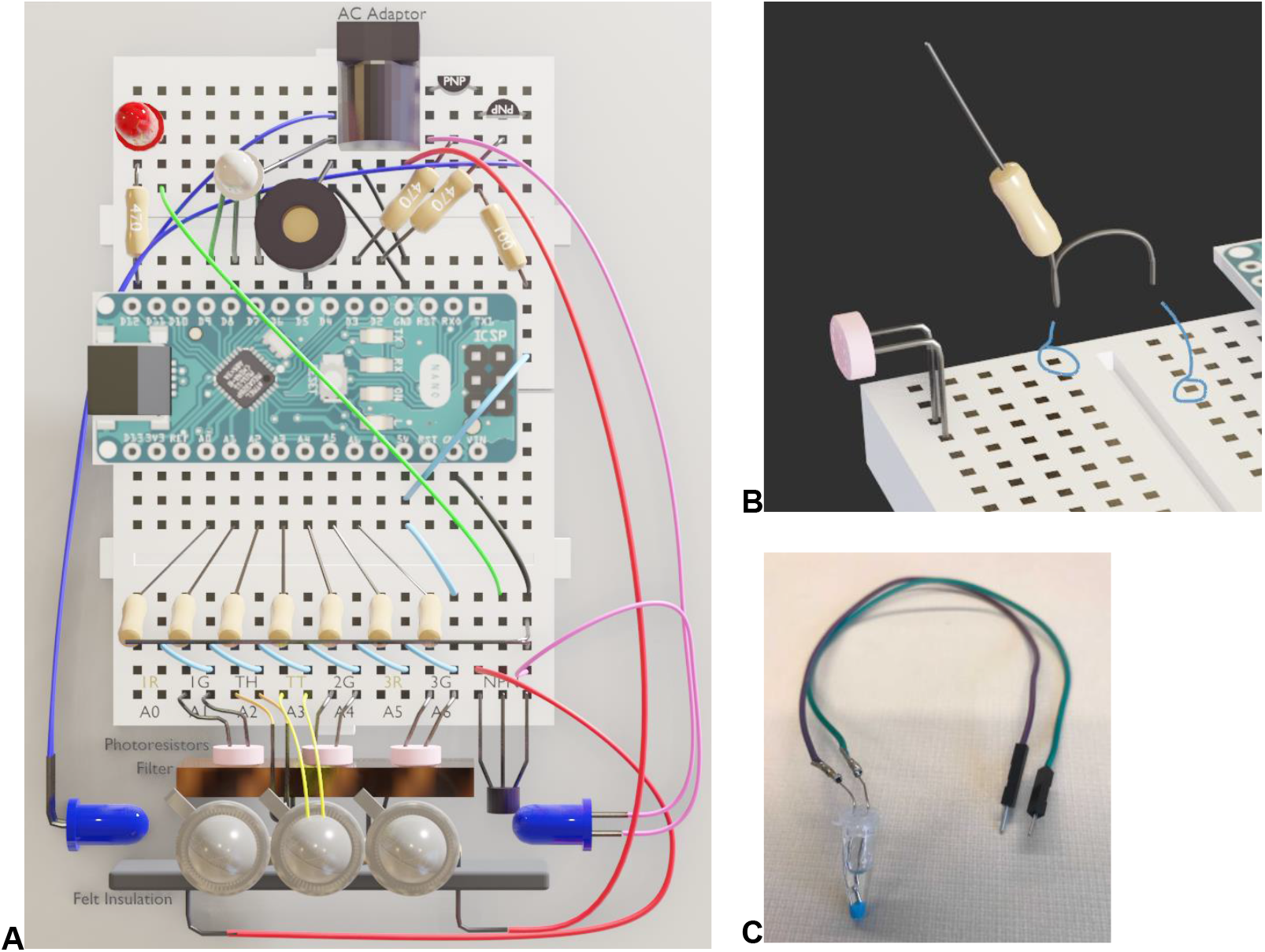
Graphical rendering of the electronics. **A**. An overhead view of the electronics. The controller (Arduino Nano) is centrally located spanning the two 170 tie breadboards. The top of the image depicts the back of the reader assembly. This area houses the status indicators such as the status LED (white with leads to D6, D7, D8) and buzzer (black ring to D5). This section also contains the heating indicator (red LED), light controls (PNPs), and AC adaptor. A 470-ohm resistor connects digital pin D12 to the red LED, which is then jumped down to the base of the NPN. The lighting control components are PNP transistors whose base is connected to digital pins D2 and D3 by 470-ohm resistors on the upper right along with their collectors to jumper wires connecting to the LED lights. A 100-ohm resistor paired with a jumper wire provides 5-volt power from the controller to the PNPs’ common emitters. The bottom of the image displays the front of the electronics featuring the sensor array that attaches to the tube holder (assembly shown in **Figure 4**). Sensors are powered by the controller’s 5-volt supply, which vary their resistance for either light or temperature, then go to analog pins A0 through A6 and have a parallel path through a 10K-ohm resistor to ground. The ground and 5-volt rails are extended to the outer board via the jumper wires shown. NPN transistor is also depicted with its leads: Collector, Base, Emitter from left to right. **B**. Single resistor bridge depiction. Folding of the resistor lead enables it to be used as a bridge to connect the sensor rail to the proper analog pin of the controller. Forceps or fine nose pliers can be used to create the fold. **C**. In-Tube thermistor.

#### Electronics: Arduino Nano Controller and Breadboards

The base of the electronics build is an Arduino nano and a pair of 170 tie solderless breadboards. Solderless breadboards were chosen to simplify reader construction and allow for flexibility with regard to component reconfiguration. However, without an enclosure, leads may become dislodged during handling of the reader. The Arduino nano is positioned to span the two breadboards. The adhesive pad of the breadboards can be used to affix the electronics assembly to the base of the housing following the completion of the electronics buildout.

#### Electronics: Sensor Array Assembly

The assembly of the sensor array begins with seven 10k Ohm resistors (R0-R6). A resistor should be placed to correspond to analog pins A0-A6 by using one of the resistor leads as a bridge (see **Figure 2B**). The second lead of the resistors R0-R5 are twisted together with the lead for R6 which is placed into the ground tie on the far right of the breadboard. Ensure that a space is left on the breadboard between each resistor. Next, insert jumper wires (or male-to-male ribbon cables) to connect each pin position to the 5-volt power supply provided by the controller. A method for this wiring can be seen in **Figure 2A**. Jumper wires are preferred over ribbon cables as they reduce the height making cable management and assembly within the housing easier. Additional jumper wires are needed to establish a 5 volt and ground rail on the portion of the breadboard opposite the Arduino nano (**Figure 2a**). The sensors and NPN are added to the breadboard following their insertion into the tube holder and heating assembly (**Figure 4**).

**Figure 3.**
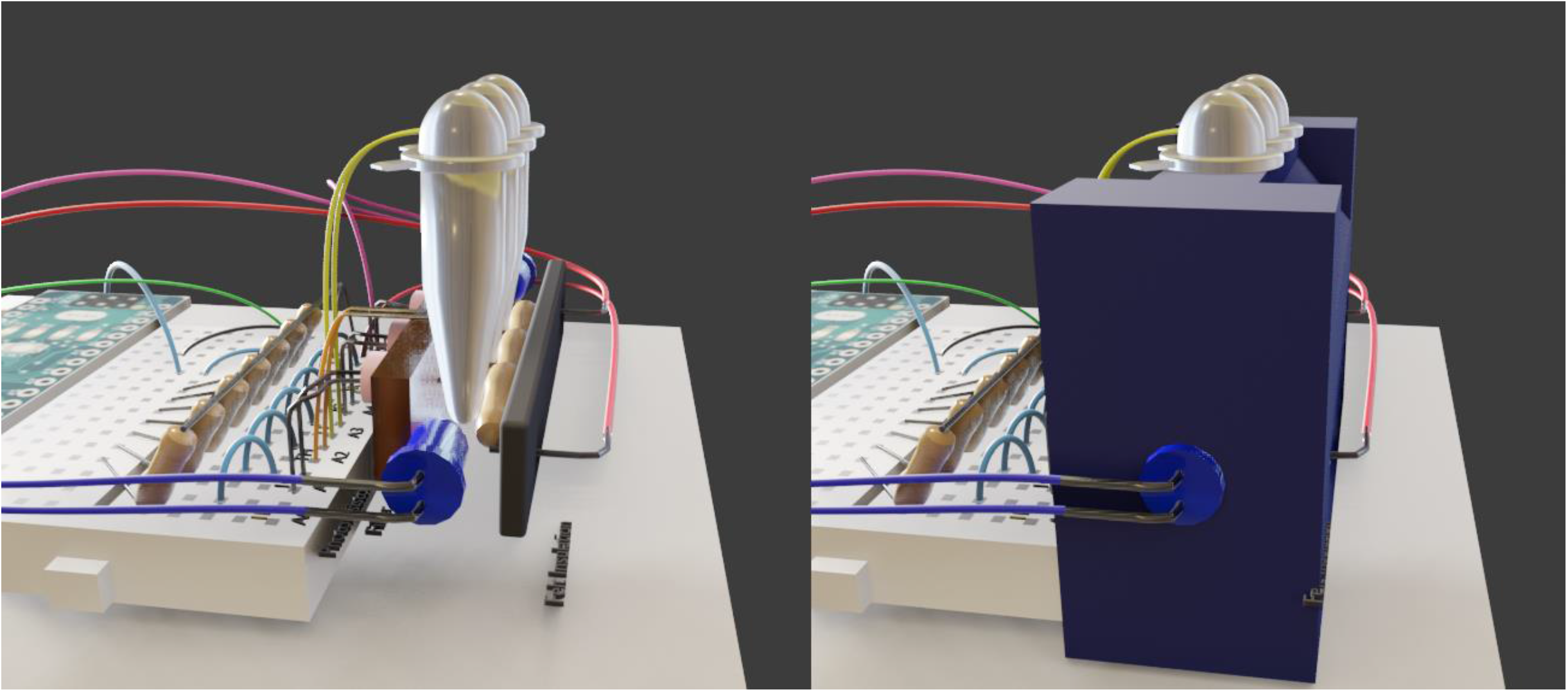
A side view of the components inside the “tube holder”. Contained within the tube holder is the heating element, in-holder thermistor, in tube thermistor, blue LEDs, and photoresistors. Clips are present to hold both the filter and heating element insulation in place. Additionally, a blue emission filter is placed in front of the photoresistors.

**Figure 4.**
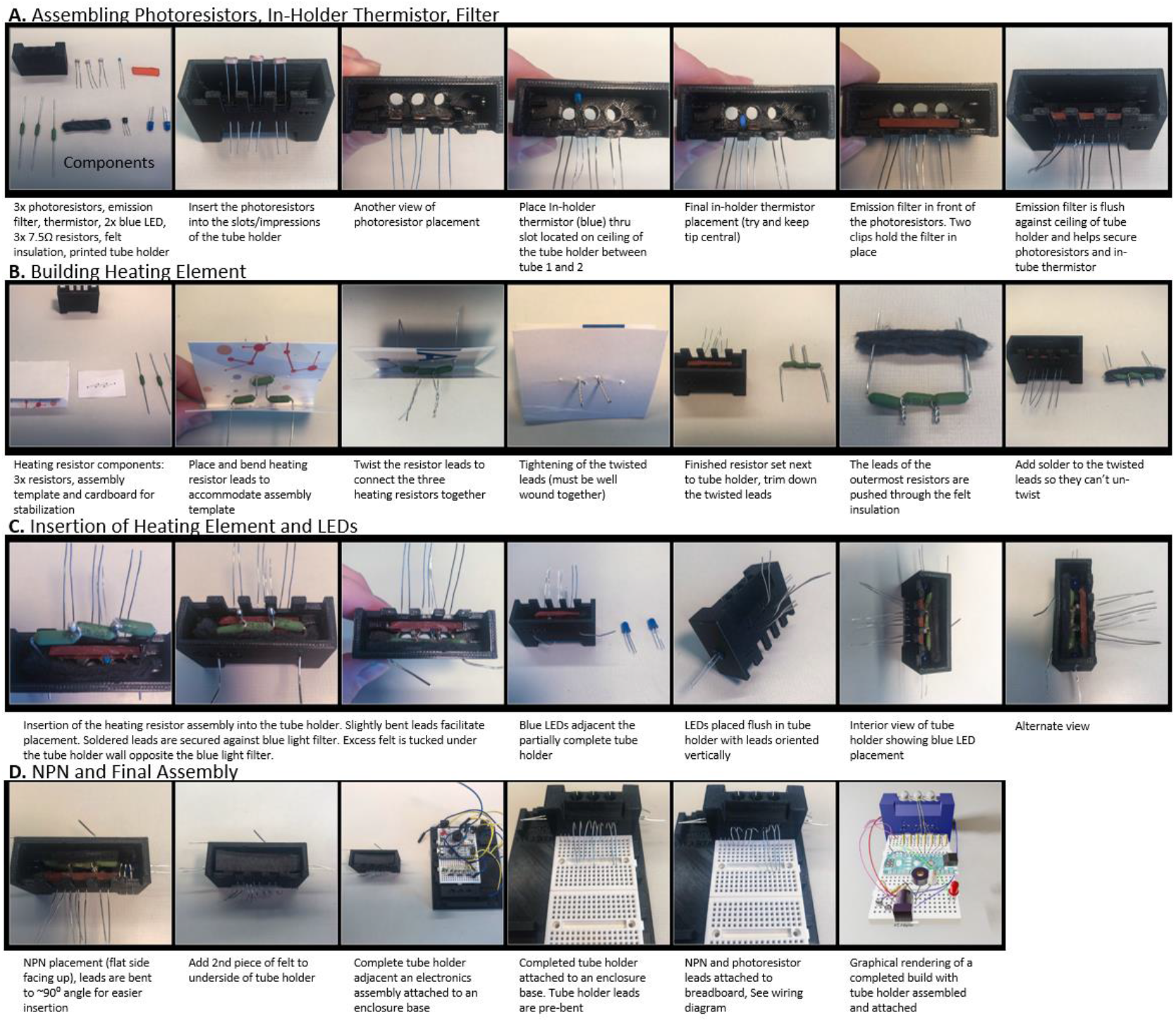
Instructions for Tube Holder construction. **A** Photoresistors, thermistor, and emission filter insertion. **B** Heating element construction. **C** Heating element insertion to the holder. **D** NPN and final assembly.

#### Electronics: Lighting Circuit Assembly

The lighting circuit assembly starts with the placement of two 470 Ohm resistors bridging the D2 and D3 rails of the controller to the outer portion of the breadboard. Each resistor should correspond to the rail of the middle position of each of the overlapping PNP transistors at the rear of the board. The leads of these resistors can be shortened to bring them lower to the breadboard. A jumper wire, coupled with a 100 Ohm resistor, supplies the 5-volt power. The placement of the cable lines up with the shared positive lead of the overlapping PNP transistors seen in **Figure 2A**. Four male-to-female ribbon cables are needed to connect LED lights to the breadboard. The male ends of two of the ribbon cables are inserted directly into the breadboard each on the non-overlapping outside ends of the PNP transistors. The female end of the cable will go to the positive lead of the LED (longer lead). The other two male-to-female cables will be used to ground each LED. The male end of the cable can be placed in any of the ground positions provided by the controller. The female ends are attached to the negative lead of each LED.

#### Electronics: Status Indicators

An optional but useful feature of the electronics are the components that constitute the status indicators. These two components are a multicolor LED and a piezo buzzer. The buzzer is placed on the D5 controller on the controller and grounded using the ground rail supplied by the controller and a jumper wire. The multicolor LED has the three short leads placed in D6, D7, and D8 and is similarly grounded. The schematic layout can be found on the top of **Figure 2A**.

#### Electronics: In-Tube Thermistor

The in-tube thermistor is used to regulate temperature in the reaction tubes. It is placed in the T2 position of the tube holder. Assembly consists of one thermistor, two male-female jumper ribbons, oil/water, one 0.2mL hinged PCR tube, and 3/32” heat shrink wrap (optional). Begin by piercing the lid of the 0.2mL hinged PCR tube to feed the leads of the thermistor. Thermistor leads are fed through the lid, so the thermistor is seated at the bottom of the PCR tube (**Figure 2C**). If using heat shrink cut two 20mm sections and cover the leads of the thermistor not inserted into the PCR tube. Trim away the plastic shielding on both the female ends of the ribbon cables using wire strippers. Insert the leads of the thermistor into the remaining female connectors. Solder is applied to the insertion point to secure the thermistor to the ribbon cable. After the solder has cooled, if heat shrink was added, cover the remaining exposed leads, and shrink using the soldering iron. Fill the PCR tube with water or oil. The 2 male leads of the jumper ribbons are attached to the breadboard in the A3 rail of the breadboard position (**Figure 2A, Figure 4**).

#### Electronics: Heating Element

The heating element of the reader went through several design iterations to find a cheap and easy-to-assemble element that could reach temperatures of 65°C within 2 minutes and to remain as low wattage as possible. Early testing using nichrome-based heating pads or small metal heating elements at the base of the tube were not sufficient to heat the reaction. This led to use of wirewound heating resistors positioned proximal to the reaction tubes. Specifically, three 7.5 Ohm 1-watt wirewound resistors connected in series. Each resistor is lined up to heat a single tube that is inserted into the reader. The control of the heating element is through an NPN transistor regulated by digital pin D12 of the controller. A 470-ohm resistor from this pin connects to a red LED, used to indicate the heater status, and a jumper wire to the base of an NPN transistor serves as the switch to turn the heat on and off. The wirewound resistor array is placed within the tube holder backed by insulation. If AC power is selected for the heating element take extra precaution by only testing heat when the reader is fully assembled. A graphical view of the heating element can be found in **Figure 3**.

#### Reader Housing Assembly

The housing of the reader was designed to consist of a minimal parts list and be printed using enthusiast-grade 3D printers. The overall design adopts a rectangular “clamshell” approach meaning that the reader is assembled on a base and the enclosure and lid of the housing clips into the base. Designs were done in the open source software Blender (http://blender.org) and were exported to STL format for printing, source files are made available in the supplement. Testing can be done without the enclosure so that most of the build is easily accessible (only plug in the USB, not the AC power until the enclosure is fitted). The housing consists of 4 main components: the base, enclosure, lid, and tube holder.

#### Reader Housing: Tube Holder

The tube holder houses the heating element, excitation LEDs, thermistors, and photoresistors. This design accommodates a three-tube compatible reader, but the third (T2, middle) tube is utilized to regulate heating. The first step of assembly is the insertion of the photoresistors and thermistors. Three photoresistors are inserted on the interior of the assembly and the leads are fed through the printed holes corresponding to each spot (**Figure 4A**). A circular indentation allows the photoresistor to be seated in the tube holder to prevent slipping of the sensor and ensure uniform positioning of all three photoresistors. A similar printed indentation is located on top of the tube holder for the in-holder thermistor. Each resistor corresponds to an individual tube slot in the holder. Following insertion of the photoresistors, the blue light filter (which is orange in color) is slid into clips on the interior of the holder. Specifications for the size of the filter can be found in **Table 3**. The heating element (**Figure 4B**) is placed opposite the filter and photoresistors. The outer leads of the heating element resistors are fed through the diagonal holes of the tube holder (**Figure 4C**). The blue LED lights are inserted into the printed ports on either side of tube holder (**Figure 4C**). Insulation is necessary between the heating resistors and the interior wall of the tube holder. Felt sufficiently insulated and prevented softening or melting of the 3D printed components while reducing heat loss. The NPN transistor is the final component inserted into the tube holder (**Figure 4D**). Following assembly, the leads of the thermistor, photoresistors, and NPN can be placed into their designated pin positions on the breadboard (**Figure 4D**).

#### Reader Housing: Base and Enclosure

The base of the reader housing serves as a platform for all of the internal components. The completed breadboard and controller are centrally located on the base. The adhesive pad found on the underside of most 170 tie breadboards can be used to affix the board and controller to the reader base. The sensor array portion of the board faces the pedestal end of the reader base. The pedestal serves as the platform for the tube holder assembly. 4 clips on the front and back of the base serve to secure the enclosure. An additional battery slot is located adjacent to the breadboard position if battery power is selected to run the reader. The enclosure serves to cover the electronics and secure the tube holder and base of the reader. Power supply and USB ports are located on the rear and side of the enclosure respectively to accommodate both power options. The final element of the enclosure is the slot for the lid which serves to cover the tube holder when in use (**Figure 5**).

**Figure 5.**
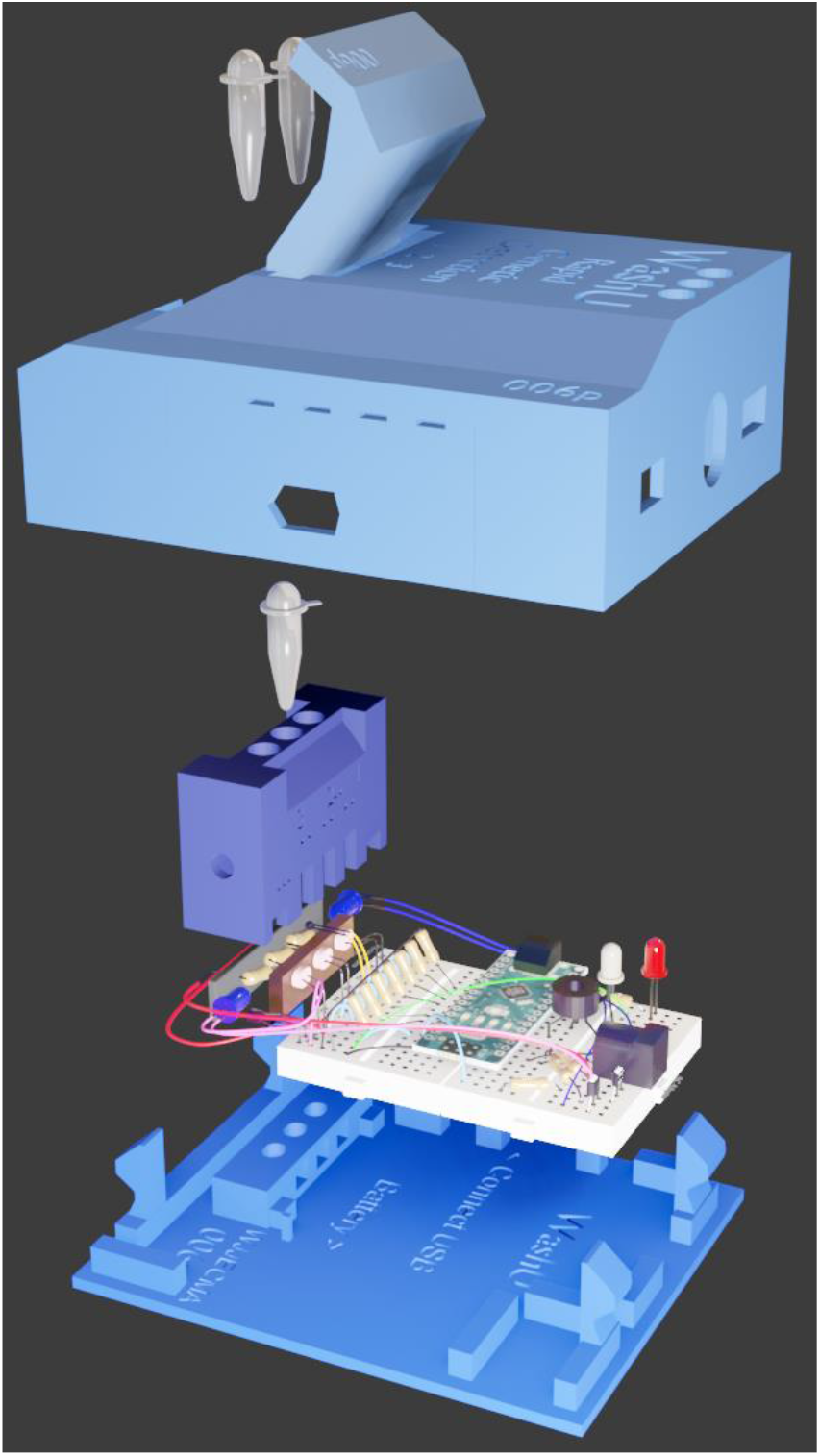
View of all components. From the bottom up, you can visualize the pieces and how they fit together. Bottom is the “base” 3-D printed part, then the electronics boards. Then the tube holder, then tube thermistor, then the “enclosure”, then the reaction tubes, then the lid. This ordering is not precise to the build, since the tube holder is assembled before all the connections to the electronics. The middle tube is separated from the outer tubes since this tube is used for the temperature control (in-tube thermistor) and is placed and tested before the top enclosure is fitted.

#### Reader Power Considerations

To increase the breadth of usage, two primary modes of power were assessed in the development of this platform. The first was a standard 9-volt battery offering the ability to use the device in the absence of an outlet and flexible deployment. Performance under battery power yielded effective testing, however, the drain on the battery limited the use of the system to 2-3 hours of continuous usage. Battery failure resulted in a loss in temperature invalidating the results of the most recent test. Following feedback from healthcare providers and analysis of different usage scenarios, AC adaptor compatibility was added to the reader. A 9-volt AC adaptor provided the ability to use the reader continuously throughout the day without the need to monitor power. The drawback of this method is the need for a 9-volt power supply and the additional safety considerations when using AC power (never plug in without the full enclosure fitted).

#### Firmware and Software

All firmware for the reader was uploaded to the Arduino nano ATMega through the Arduino IDE (https://www.Arduino.cc/en/main/software). The firmware was also developed using the Arduino IDE (C, C++). The PC software application was written in DOTNET framework. Detailed instructions on the usage of the software can be found in the protocols section. All software and firmware source code are available (https://gitlab.com/buchserlab/rgd).

### Quality Control Protocols

#### Quality Control Protocols: Initial Startup

A reader must be quality controlled following completion of the electronics build. Before starting, ensure that both the Arduino IDE (https://www.arduino.cc/en/Main/software) and dotNET Core (https://dotnet.microsoft.com/download) are installed on the PC that will be connected to the reader. Following installation, connect the reader to the PC through the USB cable provided in the kit. Upload the firmware to the device through the Arduino IDE (ensure that that proper port is selected in the tools menu of the IDE). The upload will trigger a series of light flashes from the Arduino nano if successful. Run *RGD_Control*.*exe* on the PC to start the software, which should automatically detect and connect to the reader upon startup. Navigate to the ***Settings*** tab (**Figure 7A**) to begin configuring the reader. Click the ***more*** button to open the pin assignment menu. The controller’s pins are correctly assigned by clicking the ***default*** button. Confirm that the pins are correct. The correct assignments are as follows: ST1 (A)=1, ST2 (A)=4, ST3 (A)=6, LED T1 (D)=2, LED T3 (D)=3, Thermo1 in tube thermistor (A)=3, Thermo2 in holder thermistor (A)=2, Heater (D) = 12, Buzzer (D)=5. Pin positions can be manually edited if an alternative layout is desired. The ***Push Pins (Status)*** and **Push Pins (up to Heater)** buttons are clicked once all of the pins are set correctly. The buzzer can be tested using the ***Try Buzzer*** button. Each press will cycle through the 4 different buzzer sounds. The ***Status Lights*** button confirms proper pin designation and breadboard insertion of the multicolor LED. Each press will cycle through the flashing status light LED colors (Red, Green, Blue, Cyan, Magenta, Yellow). Returning to the ***Settings*** tab, select the ***lights*** checkbox and click ***Update***. The Blue LEDs on each side of the tube holder should flash blue at rapid intervals. Deselect the ***lights*** check box and then click ***Update*** again which will turn LEDs off. Blue LEDs will flash in series with a cycle length of 2.5 seconds, and each light comes on for 500 msec once a test run is started. The timing of the LED lights and photoresistor reading frequency is outlined in **Figure 6**. A flashlight can be used to test photosensor 1 and photosensor 3 by waving the light over corresponding tube holders. The plots for Tube 1 and Tube 3 should increase when the light is above the corresponding tube position.

**Figure 6.**
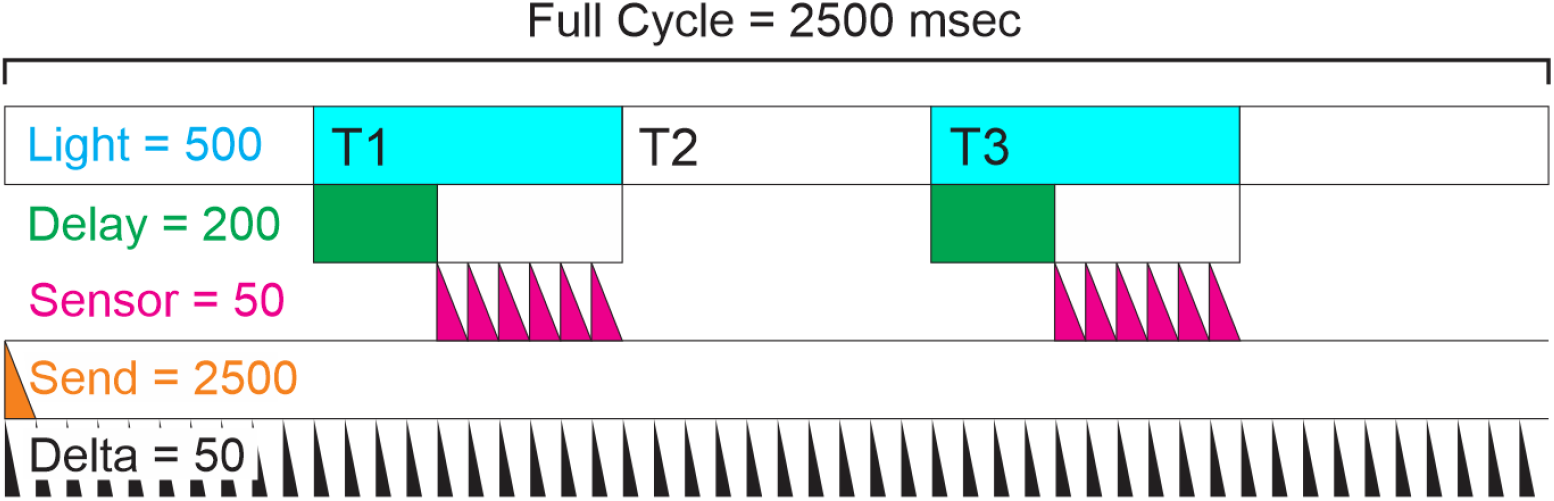
Timing of the system. The software controls the timing of the firmware through 6 different variables. Shown is a snapshot of what occurs across a single cycle (which repeats throughout the 30-minute run). The cycle length is 2.5 seconds, and each light comes on for 500 msec. The figure above shows that the Tube 1 LED comes on from 500-999 msec, then turns off, while the Tube 3 LED comes on from 2000-2499 msec. After the LED turns on, there is a delay of 200 ms before the sensors start recording the light intensity. The sensor then captures a measurement every 50 msec (which they do ∼6 times while the light is still on). These data points are averaged together and transmitted once every cycle (the ‘send’ mark). Finally, the delta parameter just sets the minimum internal delay inside the ATMega. Nothing can happen faster than this value.

**Figure 7.**
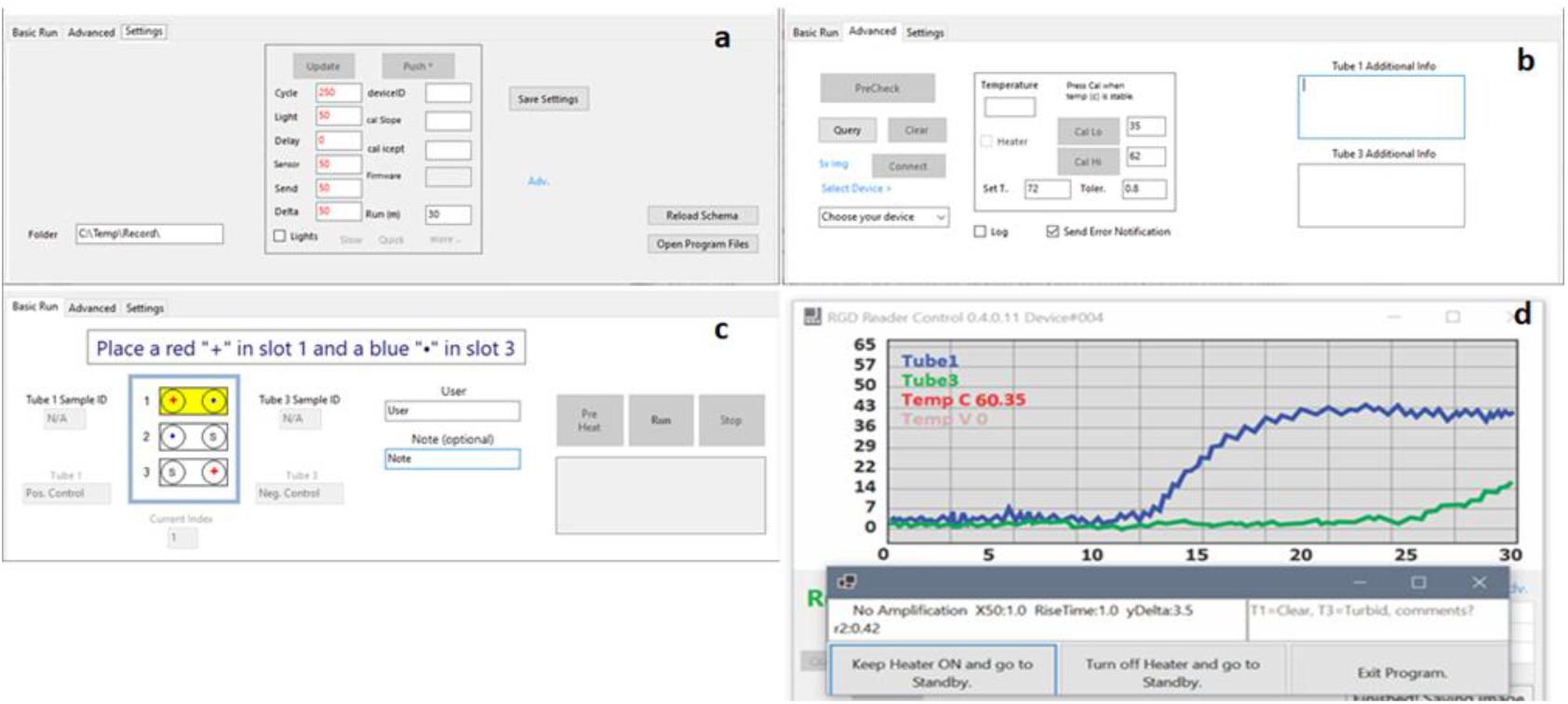
Software. **A**. Settings tab of the reader software. This tab contains the device ID information as well as calibration settings and pin designations. The destination of the run data is designated here. **B**. Advanced tab of the software. The advanced tab contains thermistor calibration inputs as well as device selection and metadata input. **C**. “Basic run” tab of the software. Following quality control all testing will be facilitated here through following the software guided instructions. Additional information such as the sample ID, user, and notes can also be recorded here. **D. “**Post run” screen displaying final amplification curves, test result interpretation, and options.

#### Quality Control Protocols: Thermistor Calibration

Begin reader thermistor calibration by removing the in-tube thermistor from tube holder position 2. Place the in-tube thermistor on the benchtop so that it is immobile. Once the TEMP C reading has stabilized record the room’s temperature in the ***Cal Lo*** section of the ***Advanced*** tab. A laser-based thermometer can increase calibration accuracy by obtaining a reading directly from the thermistor tube. Press the ***Cal Lo*** button to record this reading. Using a thermocycler (Or device capable of holding constant heat up to 60C), set the temperature to 60C and allow it to get to temperature with the in-tube thermistor. A quick rise in the displayed Temp C should become apparent. Allow Temp C to stabilize with the in-tube thermistor in the thermocycler. Enter “60” into ***Cal Hi*** section and then Press ***Cal Hi***. Navigate to the settings tab. Enter any number into the ***deviceID*** and then click ***Push*. deviceID*** is a user-assigned number to distinguish between multiple readers. A message will appear regarding the maximum number of pushes the controller can accommodate. Accept this message to complete the push. The in-tube thermistor has completed calibration once the above steps are completed.

#### Quality Control Protocols: Heating Verification

Upon completion of thermistor calibration, the heating element can be properly tested. Start by navigating to the ***Advanced*** tab and check the ***heater*** box to begin heating (**Figure 7B**). The red LED on the reader should illuminate following checking the box. As heating continues, a rise in TEMP C (displayed in the plot), should be observed. Allow TEMP C to reach 60C to ensure that the heating element is able to deliver sufficient heat. Once the set temp has been reached, examine the red heating LED. The LED should oscillate on and off depending on the set tolerance. The default tolerance is 0.8C. The in-tube thermistor should be examined to confirm that the thermistor is seated at the base of the PCR tube. This is needed to obtain the most accurate temperature reading for best reaction results.

#### Quality Control Protocols: Precheck Procedure

Navigate to the ***Advanced*** tab of the software and click ***Precheck*** (**Figure 7B**). This initiates an automatic reader self-test. The first sequence of the precheck evaluates the blue LED lights. The reader records “dark mode” data by turning off the LED lights and recording from both the T1 and T3 sensors. A second reading with the LED lights on is then taken for both sensors. The difference in sensor readings will be calculated and the delta displayed in the software. A delta of **10% or more** is satisfactory to continue. The heater is then turned on and the rise following 20 seconds is recorded. A temperature rise of at least 5 degrees should be observed. A report will be generated following the pre check and saved in the directory specified in the ***folder*** box in the advanced tab. If one of the tests fails, check the wiring for that part of the circuit.

#### Quality Control Protocols: Viral Particle Testing

Following completion of the aforementioned quality control measures we validated our readers through testing using viral particles (NIH: SARS-Related Coronavirus 2, Isolate USA-WA1/2020, Heat Inactivated, NR-52286). Testing using viral particles in clinical matrix (saliva) was performed to validate the chemistry as well as reader performance in a simulated test scenario. During this testing, software instructions were followed to ensure appropriate controls were run. The order of the controls is displayed in **Figure 7C**. A red “plus” corresponds to a positive control and a blue “dot” corresponds to a negative control. The sample condition “s” during this process represents the viral particle testing. Testing was repeated four times per reader using the same number of viral particles and reaction chemistry lot. Appropriately dispose of the reaction tubes following the completion of a test and take care to not open the reaction tube.

Extensive internal testing using this paradigm allowed the validation of multiple readers and batches of reaction chemistry. If appropriate facilities are available, viral particle testing can be undergone, however, it is sufficient to replace this testing with the non-biohazardous gene block controls spiked into clinical matrix (saliva).

### Testing Protocol

#### Testing Protocol: Sample Testing

All sample testing is conducted through the **Basic Run** tab of the software (**Figure 7C**). Upon startup, the reader should automatically be detected (ensure that the reader is plugged into a compatible USB port). Press the ***PreHeat*** button to begin heating the reader. Samples cannot be run until the reader has reached the set temp of 60C. The software will instruct the user to perform a set of initial runs using controls to ensure proper reader functionality. These runs consist of a positive and negative control. Ensure that both control tubes are fully thawed before being placed into the reader. The user will be notified if the run has failed and that sample testing should not proceed prior to examination of the reader and chemistry. Following each quality control run and sample test the user will be provided with a screen displaying the amplification curve, results, and options for continuing (**Figure 7D**). To continue running samples select the *Keep Heater ON and Go to Standby* option. The other options will either turn off the heater or prompt the user to proceed with the shutdown QC run.

Following the initial quality control runs, the software indicates what tube position the reaction tubes should be placed as well as what reaction tubes are needed. The appropriate sample and control tubes should be removed from the freezer and thawed together. Do not place the control tube in the reader before the sample tube. Remove a sterile loop from its packaging and touch the loop either to the surface of the tongue or beneath the tongue to collect the saliva needed for the test (no scraping). Insert the loop into the sample tube, discard the loop, and close the sample tube. Place both tubes in the reader, close the lid, and press run. The duration of the run is 30 minutes. Allow the run to complete before interpreting the results. The software will perform an analysis upon completion of the run and determine if there was amplification in the sample (called “product amplification”) as well as other parameters of the curve fit (**Figure 7d**). After the last sample, the software will instruct one more QC run, and then shutdown.

## Results

### Analytical Limit of Detection in Simulated Clinical Matrix

In order to determine an analytical limit of detection (LOD), heat inactivated viral particles were spiked into a simulated clinical matrix. This simulated matrix consisted of the 40 µl LAMP reaction with 1 µl of saliva (from 5 different individuals) and a known number of viral particles (NIH: SARS-Related Coronavirus 2, Isolate USA-WA1/2020, Heat Inactivated, NR-52286). Viral titers of 10^4 were used to quality control readers prior to LOD testing (with 100% success). Testing consisted of viral particles levels of 100, 30, and 10 in each reaction, aka per 1 µl of saliva (**Figure 10**). Viral loads down to 100 viral particles (VP) per µl of saliva were reliably detected in 95% of runs. 30 viral particles were detectable at a rate of 92%. Given recent research into viral loads showing less than ∼100 copies per microliter were not infectious (La Scola et al., 2020; Quicke et al., 2020; Wölfel et al., 2020) an LOD near 30 particles is sufficient to detect infectious levels of SARS-CoV-2.

**Figure 10.**
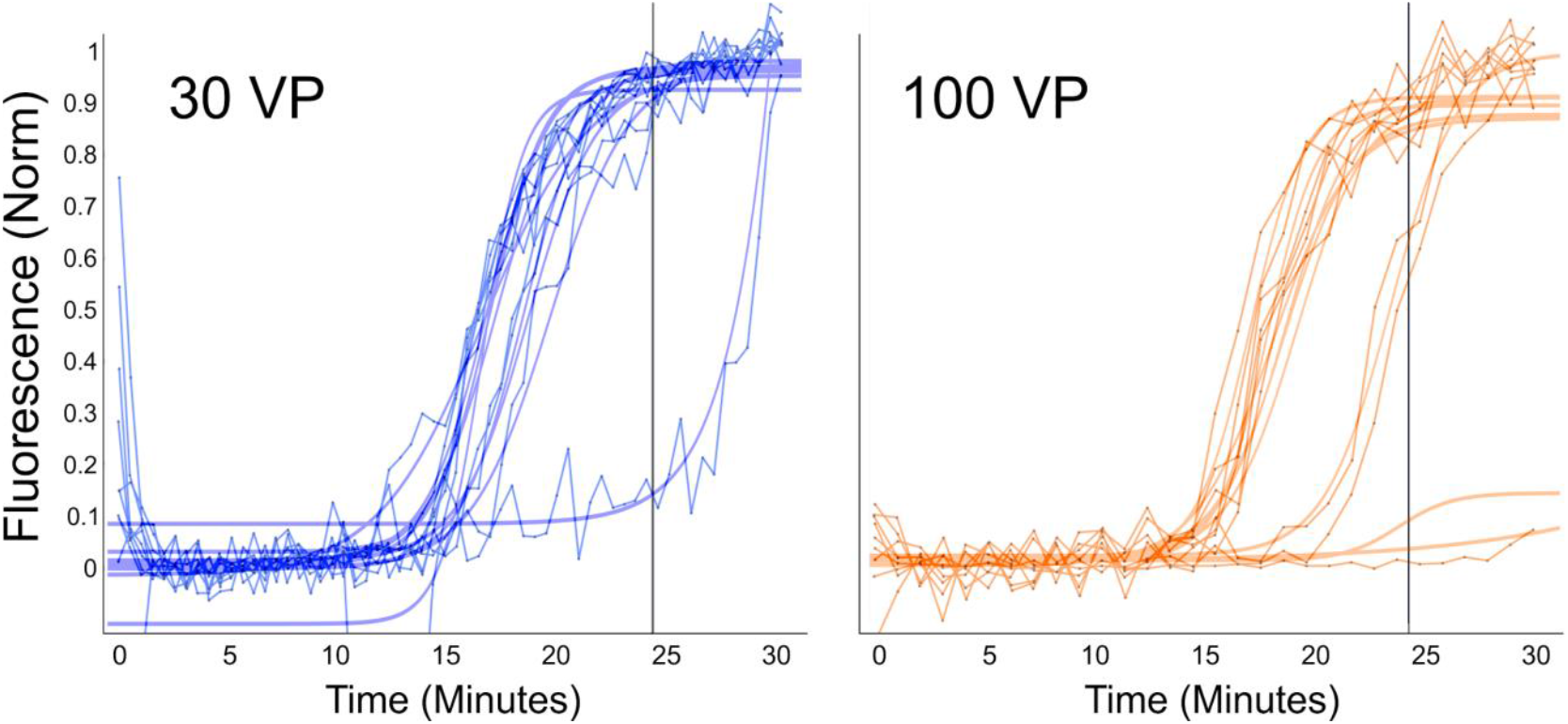
Limit of Detection using Viral Particles in Clinical Matrix. Known concentrations of viral particles were spiked into reactions with one microliter of loop-delivered saliva. Readers used to evaluate the LOD had previously passed initial quality control to eliminate reader-based reaction failures. A LOD of between 30 (92%) and 100 (95%) viral particles was determined after repeated testing with decreasing concentrations. 10 viral particles (not shown) was able to be detected at a rate of 85%. The traces are directly saved from reader (as constructed per the instructions above). The raw trace and a curve fit are displayed for each replicate.

### Clinical Saliva Results

A series of clinical samples were run on several readers to determine performance of the test using saliva samples collected from patients with corresponding RT-qPCR NP swab results. All saliva sample collection was approved by the institutional review board at Washington University School of Medicine (WU350, IRB#202003085). Informed consent was obtained for all samples. All of these saliva samples were also run with the Fluidigm Advanta Dx SARS-CoV-2 RT-PCR Assay testing system. A total of 117 samples were run blinded to the NP swab result. As in all testing done with this system, each reader started with QC, and controls were placed on the reader as instructed by the software throughout the experiment. All analytical thresholds for the determination of positive, negative, and ambiguous results were predetermined prior to testing and unblinding. A view of the amplification curves for the first 38 samples is shown in **Figure 11 A**. The red curves represent those samples called positive by NP swab while the blue shows negative samples. Overall, the sensitivity of the assay was calculated to be 87% with a specificity of 95.8%. All samples used in the trial were transferred into the tube containing the test chemistry with a one microliter inoculating loop unless insufficient sample was present in which case a pipette was used to transfer the sample.

**Figure 11.**
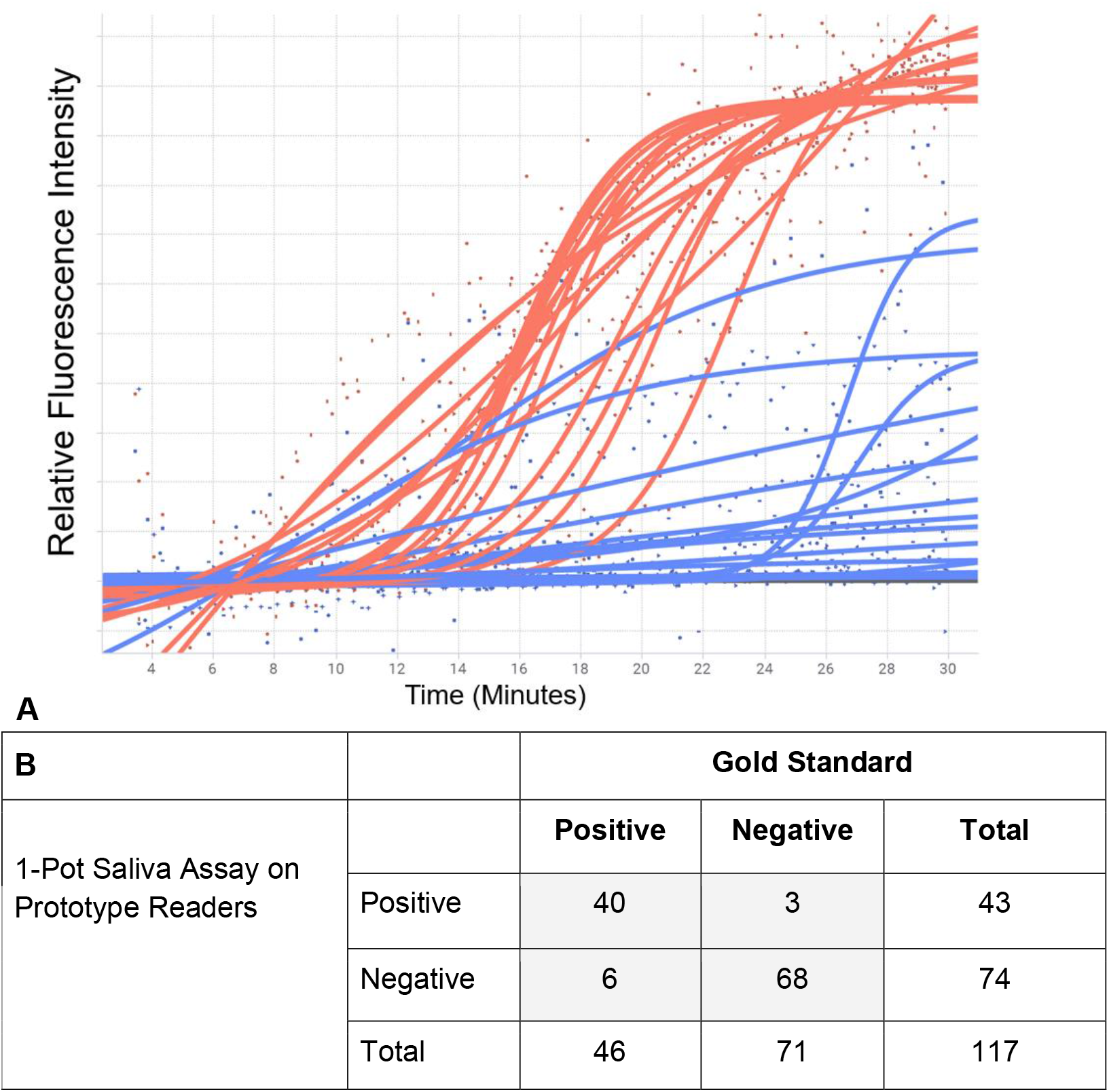
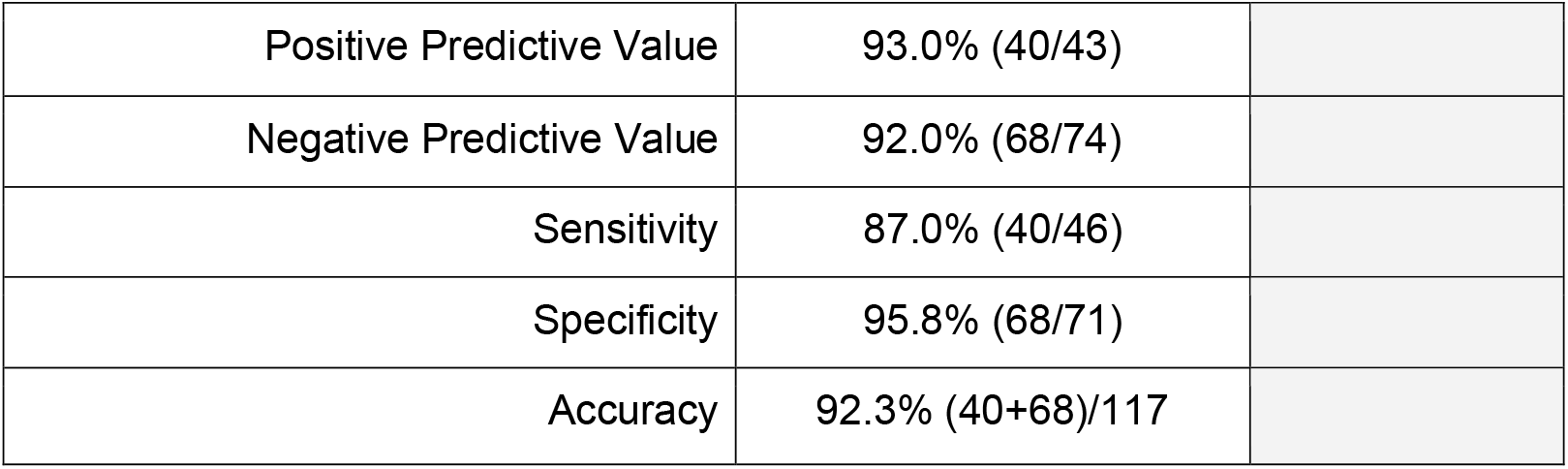
Results for POC readers with 1-color 1-pot LAMP reactions. **A**. Amplification results of 38 blinded clinical samples run with pipetting to the reaction tube. **B**. Full results from 117 total subject’s saliva, all blinded. The gold standard comparator used in this analysis are results from the Fluidigm saliva assay as well as NP swab RT-qPCR results when available.

## Discussion

The current reporting delays and supply chain limitations in COVID-19 diagnostics experienced by centralized laboratory testing platforms demonstrates the need for distributed, accessible, and more rapid testing solutions. Antigen tests provide one alternative to meet these criteria, however, while offering both a cost savings and turnaround speed advantage, they suffer from lower sensitivity and specificity. Here we present a distributed extraction-free saliva-based molecular testing platform with a less than 30-minute run time. This solution consists of an isothermal LAMP reaction coupled with a low cost, heated fluorescence reader. The results of this are a one-pot, genetic, real time, point of care platform technology for pathogen detection.

The development of this reader was driven by the idea of creating a platform technology that could be readily and rapidly deployed across a wide range of use cases. As a result, the reader only occupies a small footprint and is compatible with both AC and battery power. Battery power allows the user to perform field testing and deploy the solution in under-resourced environments. The AC power option allows for repetitive testing in areas where central laboratory-based testing is insufficient or unavailable. To achieve a simple reader design, the chemistry constituting the reaction needed to require little to no sample manipulation as well as minimally invasive conditions. As a result, the isothermal nature of RT-LAMP provided an adequate solution. Significant development was needed to modify RT-LAMP reactions to overcome the reaction inhibitors present in saliva. The key modifications that allow direct saliva addition are believed to be the larger reaction volume (25 to 40 µL) coupled with the additional BST 2.0 added to the reaction. Volumetric inoculating loop sample delivery is also a unique advantage of this solution, as it allows for sample self-collection and eliminates several supply chain vulnerabilities of more complex sample collection methods. Under simulated conditions, the reaction can accommodate up to 3 microliters, allowing for alternatives such as disposable dental micro brushes to be used. The result is a one-pot, RT-LAMP driven platform that is easy to assemble and widely deployable. However, several drawbacks are present in the current state of this platform. Regarding the reaction chemistry, the three primary issues are nucleic acid contaminants, cold chain limitations, and single-color design. The first, contamination, was observed during the production of the reaction chemistry. The result was amplification in negative controls as well as false positive results. Additionally, post reaction positive controls have the potential to themselves be contaminants if not discarded properly. The second issue, cold chain, refers to the current constitution of the reaction chemistry. Currently, the reaction is room temperature stable for up to 4 hours, 4C stable for 2 days, and − 20C stable for several months. This limits the usage of the reaction in areas where cold chain cannot be maintained. It also adds complexity to reagent storage. A proposed solution to address this is the development of a lyophilized version of the reaction in which the reaction is reconstituted in a shelf stable buffer prior to use. This would increase the field deployability of the technology. Lastly, the one-color nature of the reaction limits the ability to determine proper sample addition. The use of a general nucleic acid binding dye in the current format reduces specificity and cannot work to ensure sample addition. A two-color system using targeted probes allows for the use of an extraction control such as RNAse P (**Figure 12**). Several approaches were evaluated in the ongoing effort to develop a two-color compatible system. The approaches evaluated included OSD (Bhadra, Riedel, Lakhotia, Tran, & Ellington, 2020), loss of fluorescence probes (Hardinge & Murray, 2019) and MERT LAMP (Wang et al., 2015). Promising results in both simulated and clinical scenarios using both loss of fluorescence and the MERT LAMP approaches have been achieved. The reader can be modified to become a two-color system through the addition of a second set of LED lights and photoresistors with an appropriate filter. The limitations of the reader primarily surround the manual assembly. The first is the ability for components to shift or become dislodged. This is true of both the breadboard, due to it being solderless, and the heating element. A solution for the breadboard issue is through using printed PCB boards instead of solderless breadboards. Additionally, a machined heat block that accommodates the lights and sensors can be used to replace the heating resistors. A prototype is currently being tested and is showing promising results. A final observation regarding the reader is the impact of electrical noise on the sensors. The signal trace observed varies depending on where the reader is used. In areas with more AC-powered equipment, a noisier signal is observed. One possible solution would be the addition of low pass filters to the electronics build to reduce the overall electrical noise.

**Figure 12.**
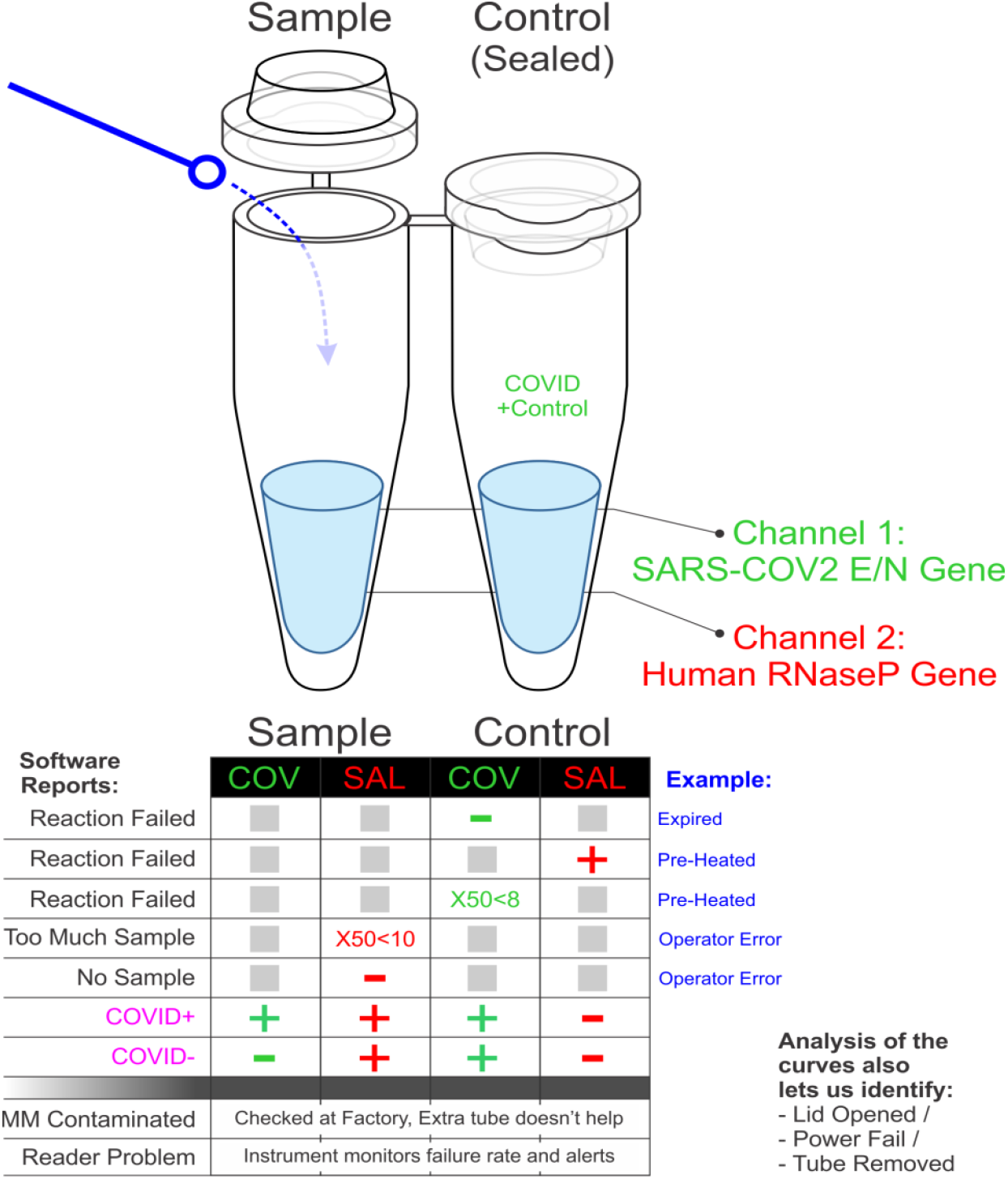
Proposed 2-color, 1-pot reaction, and interpretation schema.

The platform laid out in this manuscript provides a guide and performance evaluation of a rapid, one-color, genetic, point-of-care diagnostic detection system for SARS-CoV-2. This system is composed of a one-pot, RT-LAMP reaction from a crude saliva sample coupled with a low-cost digital reader. The results of this diagnostic are monitored real-time and interpreted automatically by the associated software. As a result, this system provides a viable option for self-testing for laboratory personnel or used in laboratories to run LAMP reactions with minimal equipment.

### On Site Trials: Emergency Room

In addition to testing these POC readers in the laboratory, we also sought to test them in the ‘field’. Initially, we placed two readers in the emergency department of BJC Hospital. There, research coordinators would collect saliva from COVID-19 suspects by placing the sterile loop under a patient’s tongue and dipping the loop into a reaction tube. The coordinator would then return to a different location with the readers, insert the tube, and run a test. Many subtle design and software changes were made based on feedback from the coordinators during this test. Two very important findings were made during this evaluation. First, we realized that since the instruments were left on for 12+ hours at a time, that the water-based thermistors would evaporate, throwing off the heating. This caused us to move to an oil-based system. We also realized that subtle shifts in the heating element resistors could cause the heating of tubes 1 or 3 to be different than the heating of the thermistor tube as well as create heating discrepancy between T1 and T3. The second in-holder thermistor was added to address that. Finally, some readers we constructed were less stable than others, and we learned that we needed to constantly run controls to ensure that everything was behaving normally. The current version of the software requires controls to be run throughout the day, ensuring proper reader functionality.

### On Site Trials: Nursing Home

We also took four readers to a local nursing home where research coordinators took samples from residents who had previously contracted, but have now recovered, from COVID-19. One of the important lessons learned at this location was that the loop-mediated saliva collection was superior for several reasons. Both a standard saliva sample (spit-in-tube), and a loop-based saliva sample were taken from the residents. Many residents could not give the standard sample, either because they were too dehydrated, or because they had trouble expectorating.

All residents were able to give saliva by the loop, and much preferred this method to the nasal swab or to spitting. The results from these 22 patients were analyzed and the readers correctly called 20/21 (95.2%) as negative. One sample was called positive by our reader, but not by nasal swab, and we were unable to obtain a larger spit sample to replicate on a Fluidigm platform. The one resident that was still positive by NP swab was not detected in this saliva test. The NP Swab result was near LOD on the Roche Cobas system, and the subject had been symptom free for several weeks.

Point of care testing for highly infectious agents with molecular accuracy remains an important feature of epidemiologic control. The proposed system elaborates a solution that is currently patent-pending and research-use-only, since it has not gone through FDA approval.

## Supporting information

Reader 3D Printing .stl Files

Reader Blender Rendering

## Data Availability

All source code and schematics associated with manuscripts have been made available

https://gitlab.com/buchserlab/rgd

## Acknowledgements

We would like to thank Catrina Fronick for all her help throughout our clinical sample testing, Seiler Instruments & Manufacturing for machining prototype heat blocks. Thanks to NHC Healthcare in Maryland Heights for allowing the IRB saliva testing and Adriana Rauseo Acevedo, the Emergency Care Research Core including, Aaron Day, Kristin Stansfield, Kate Smith, Bronson Flint, Eric Raines, Jamie Rolando, Michelle Stezovsky, Misty Tribout for coordinating sample collection. We also thank Zander Meitus, James, and Margaret Plews-Ogan for their input on design, UI, and implementation improvements. Chad Storer, Ethan Storer, and Kyle Kniepkamp performed all the 3D printing and slicing. Josh Langmade and Xuhua Chen also assisted on the many aspects of the chemistry improvements.

These studies were supported by donations to the WUSM COVID Research fund, the Department of Genetics, and the McDonnell Genome Institute. We thank Barnes-Jewish Hospital, the Institute of Clinical and Translational Sciences (ICTS), and the Tissue Procurement Core, which provided saliva samples.

---

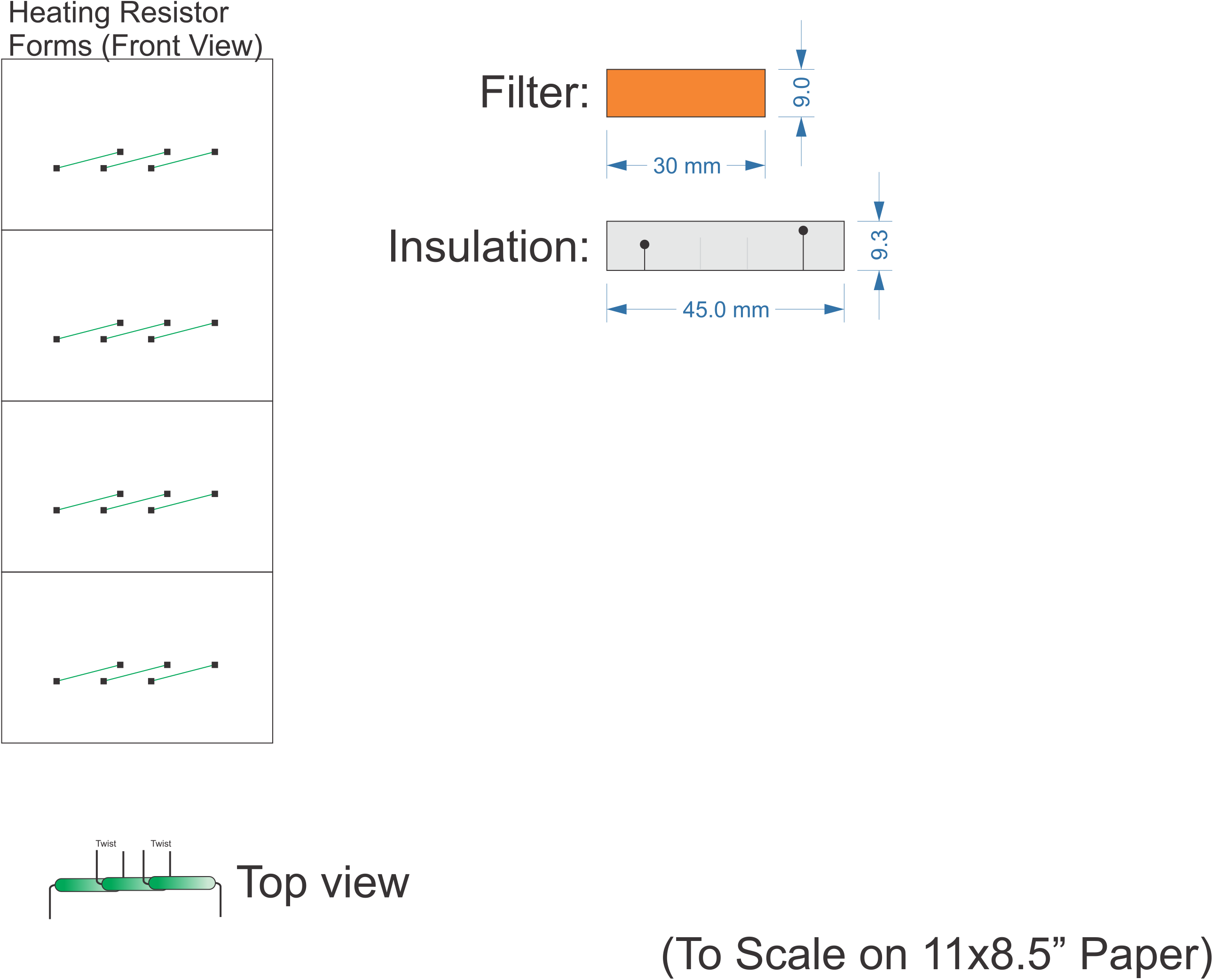

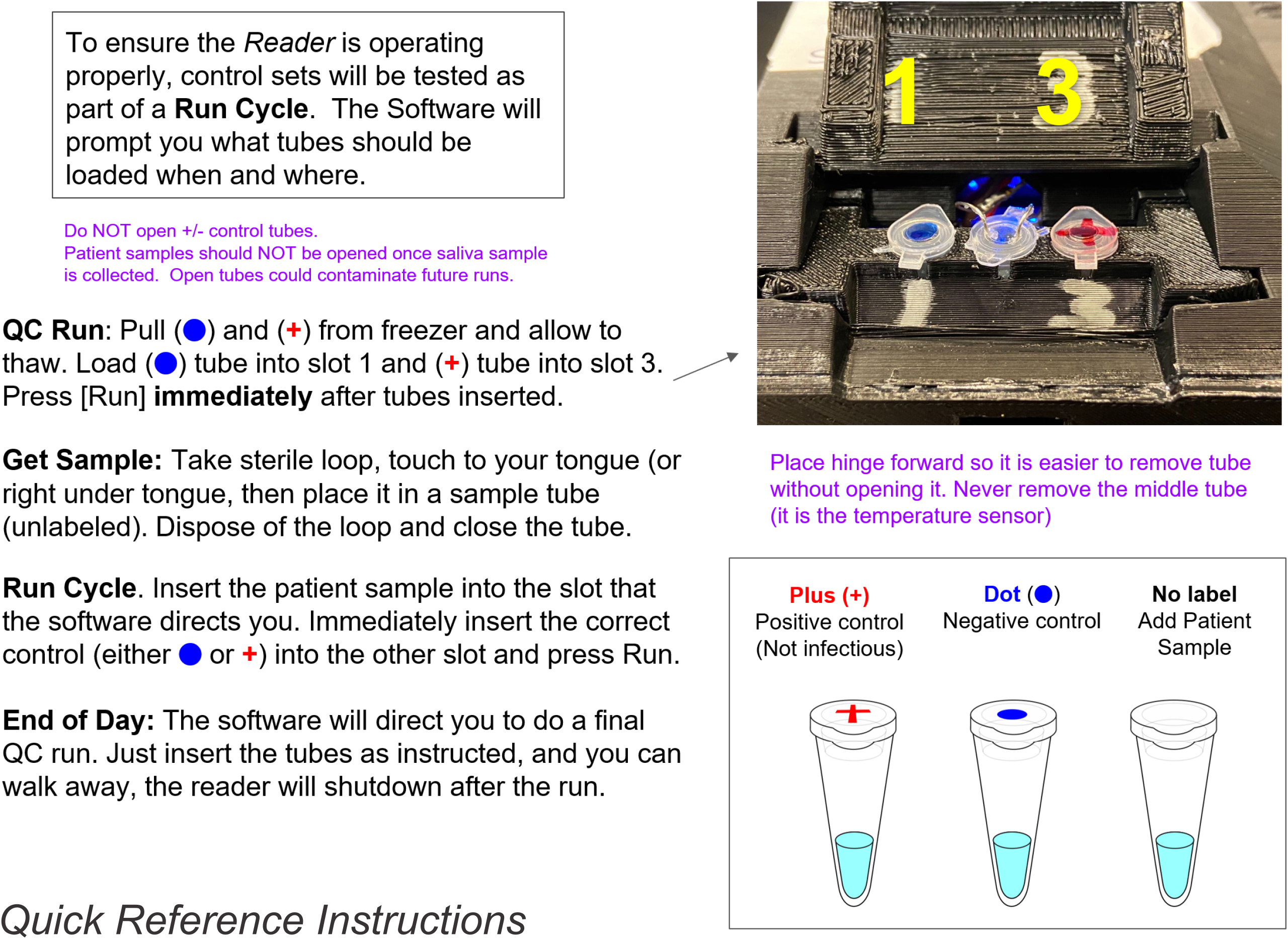

